# Accuracy and error patterns of ChatGPT-4o for real-time English–Nepali voice translation: A cross-sectional field evaluation in rural Nepal

**DOI:** 10.64898/2026.08.25.26361303

**Authors:** Alyssa Mandich, Shirsha Koirala, Sophie Westen, Saisha Adhikari, Ashraya Acharya, Abha Shrestha

**Author notes:** Corresponding author (AM).

## Abstract

Language discordance can impede community-based research and health communication where trained interpreters are limited. Although multimodal artificial intelligence systems (AI) can provide real-time spoken translation, performance with under-resourced languages during spontaneous field interactions remains poorly characterized. We evaluated ChatGPT-4o during bidirectional English–Nepali voice translation in a community setting near Dhulikhel Hospital, Nepal.

In this cross-sectional field study, 30 primarily Nepali-speaking adults were recruited by convenience sampling. ChatGPT-4o mediated conversations using standardized English questions and spontaneous Nepali responses. A bilingual Nepali–English reviewer assessed 485 translated utterances using a 3-point accuracy scale and an inductively developed framework for translation and conversational deviations.

Of 485 translations, 282 (58.1%) received the highest accuracy rating, 134 (27.6%) a moderate rating, and 69 (14.2%) the lowest. Mean accuracy was higher for English-to-Nepali than Nepali-to-English translation (2.63 ± 0.53 vs 2.23 ± 0.86); 63 of 69 low-accuracy translations (91.3%) occurred in the Nepali-to-English direction. Among 329 deviation tags, the most frequent were distortion of intended meaning (17.1%), overly formal or unnatural phrasing (14.7%), omission (14.2%), and addition of content (11.5%). Some fluent outputs substantially altered meaning or introduced information not expressed by the speaker.

ChatGPT-4o demonstrated potential for real-time English–Nepali communication but also produced errors that could alter interpretation of participant responses. Accuracy was lower and more variable for Nepali-to-English translation; however, translation direction was confounded with input type because Nepali inputs were spontaneous and English inputs standardized, limiting conclusions about directional performance. These findings support cautious use for low-stakes conversational exchange and human verification when errors could affect research validity, clinical decisions, or participant understanding. As multimodal AI evolves, performance should be reevaluated across languages, real-world conditions, and model versions, with bilingual oversight and community partnership remaining central to responsible use.

**Author Summary:** Being able to understand and be understood is essential in health care and community research. Yet trained interpreters are not always available, particularly in rural or resource-limited settings. Voice-enabled artificial intelligence could help fill some communication gaps, but a translation can sound convincing without accurately representing what a person said.

We examined how ChatGPT-4o performed during real English–Nepali conversations in a community setting in Nepal. Rather than testing carefully prepared written sentences, we studied spoken exchanges that included natural pauses, varied responses, background conditions, and culturally specific ways of speaking. We found that the tool often communicated the basic message, but it also changed meanings, omitted details, added information, and sometimes used language that sounded unnatural to Nepali speakers. Importantly, some inaccurate translations remained fluent enough that an English-speaking listener might not recognize the error.

Our study shows why natural-sounding artificial intelligence should not automatically be considered reliable. These tools may assist with selected low-risk conversations when other language support is unavailable, but they should not replace trained interpreters or bilingual review when misunderstandings could affect people’s health or representation in research.

## Introduction

Language discordance can interfere with nearly every stage of community-based research and health-related communication, including participant engagement, accurate exchange of information, understanding of community experiences and priorities, and the development of trust between individuals, researchers, and healthcare professionals [1,2]. Professional interpreters generally provide the most reliable support for communication across languages and are associated with better comprehension, satisfaction, and quality of care than ad hoc interpretation or communication without language assistance [3,4]. However, professional interpretation is not consistently available; cost, workforce limitations, insufficient integration into health systems, and geographic barriers may restrict access to qualified interpreters, particularly in rural or resource-constrained areas [1,2]. Interpretation may be especially difficult to arrange for unscheduled, time-sensitive, or community-based encounters. These limitations also pose meaningful challenges for global health research, where inaccurate or incomplete translation may misrepresent participants’ experiences, obscure community priorities and compromise the validity of collected data.

Digital translation tools have consequently attracted interest as a means of supporting communication when language-concordant staff or qualified interpreters are not immediately available [5,6]. Earlier machine-translation applications primarily used phrase-based statistical models for written text, while conventional speech-to-speech systems relied on separate speech-recognition, machine-translation, and speech-synthesis components [5,7]. In contrast, recent multimodal large language models can now process both spoken and written input and generate translated speech or text through a single conversational interface, enabling more natural interaction [8,9]. These systems are becoming increasingly accessible through consumer smartphones and computers, allowing users to initiate real-time translation without specialized equipment or advance scheduling. OpenAI’s ChatGPT has been proposed as one such tool for multilingual communication in low- and middle-income countries, where it is becoming more accessible and may offer rapid and comparatively low-cost support for field research, healthcare, and other community-based interactions [10]. However, accessibility and convenience do not establish reliability. General-purpose artificial intelligence systems were not developed or validated as substitutes for professional interpreters and may not provide the consistent accuracy and human oversight required for high-stakes communication.

Nevertheless, the unprecedented growth of artificial intelligence has prompted evaluation across a range of multilingual settings, with some studies reporting promising performance. Recent evidence suggests that large language models can produce translations that are fluent, context-sensitive, and, in some cases, more accurate than conventional machine translation systems [11–13,13]. In a comparison of English discharge instructions translated into Spanish, Chinese, and Russian, ChatGPT performed as well as or better than Google Translate at the sentence level, although complete instruction sets frequently contained at least one error [11]. Evaluations across written academic, biomedical, and general-language materials have similarly shown that generative pretrained transformer-based systems can preserve contextual meaning and generate natural, readable translations across a range of settings [11,12,14].

However, natural-sounding output may give the appearance of accuracy even when substantive errors are present. AI systems may distort intended meaning, omit relevant information, or introduce statements that were not present in the original input, creating particular risks in nuanced community-based or patient-facing interactions [15]. Research using hospital discharge notes containing deliberately inserted false recommendations found that large language models often accepted the fabricated information rather than identifying it as inaccurate, demonstrating their susceptibility to misinformation presented in formal clinical language [16]. Although translation failures in field research may not always carry the same immediate consequences as errors in clinical care, they may still produce serious misunderstandings, propagate misinformation, distort research findings, and undermine trust within participating communities. Therefore, careful evaluation of the specific errors these systems produce in real-world settings is essential. Identifying how and under what conditions failures occur may help users anticipate limitations and distinguish contexts in which AI-mediated translation may be useful from those in which professional interpretation remains necessary.

Evaluating these limitations in voice-based translation requires consideration not only of the translation model itself, but also of the conditions under which spoken language is captured and processed. Spoken input must first be detected and recognized before it can be translated and rendered as an audible response, creating multiple opportunities for error [5]. Performance may be affected by background noise, low speech volume, accents, dialects, pauses, disfluencies, mixed-language speech, and difficulty distinguishing speakers [9,17]. These challenges may be greater for low-resource languages, for which annotated speech and language-specific training data are limited [9]. Because both speech recognition and translation performance depend on the availability and quality of language-specific data, findings from well-resourced languages may not generalize to languages with more limited digital representation.

Nepali is one such language, with comparatively limited representation in digital translation and speech-recognition research. Compared with languages represented in large parallel text and speech datasets, Nepali has fewer digital translation resources and presents linguistic and cultural features that may complicate automated translation [17–19]. Nepali commonly uses subject–object–verb sentence structure, context-dependent pronouns, honorific distinctions, culturally specific expressions, and regionally variable forms [19]. Prior evaluations of Nepali machine translation have identified lexical, syntactic, semantic, and cultural errors, including difficulty preserving honorifics, idiomatic meaning, and contextual nuance [20]. Engineering studies have also described the limited availability of high-quality Nepali speech data as an obstacle to the development of robust speech-recognition and speech-translation systems [7,9,17]. These limitations underscore the need to assess whether automated English–Nepali translation preserves accuracy, contextual meaning, and culturally appropriate forms during real-time spoken interactions.

Despite growing interest in AI-mediated translation, much of the existing evidence is based on written sentences, standardized datasets, prepared medical instructions, or retrospective comparisons with reference translations [11,14,21]. Far fewer studies have examined real-time voice translation during spontaneous, bidirectional conversation under field conditions. This distinction is important because spontaneous conversation likely reflects how widely accessible AI translation tools are used in practice, where users speak directly to the system in uncontrolled environments rather than submit carefully prepared written text. Natural speech also includes hesitation, repetition, incomplete statements, culturally grounded expressions, shifts in turn-taking, and environmental noise that are largely absent from controlled written evaluations. Evidence is especially limited for spoken English–Nepali translation and for community-based interactions in rural Nepal, where a general-purpose AI system mediates communication without a professional interpreter participating in the exchange.

We therefore evaluated the feasibility and translation fidelity of ChatGPT-4o’s real-time, bidirectional English–Nepali voice translation during low-risk community-based interviews near Dhulikhel Hospital in Nepal. We assessed utterance-level translation accuracy and examined differences in performance by translation direction and interview question. Using an inductive, data-driven coding framework developed from deviations observed in the study transcripts, we also characterized the semantic, linguistic, cultural, technical, and conversational ways in which translated exchanges differed from the speakers’ intended messages. By evaluating both overall accuracy and the specific types of deviations that occurred, this study provides a focused assessment of ChatGPT-4o’s performance in real-time English–Nepali communication under community-based field conditions.

## Methods

### Study Design and Setting

This study employed a cross-sectional observational design to evaluate the feasibility of real-time Nepali–English voice translation using a large language model in a routine conversational context. The study was conducted at Dhulikhel Hospital, a community-based tertiary care institution in Kavrepalanchok District, Nepal. The project was carried out in collaboration with Dhulikhel Hospital staff as part of ongoing efforts to evaluate digital communication tools in multilingual global health contexts where professional medical translation services are not consistently available. Field data was collected in July and August of 2025.

### Participants and Recruitment

Participants were recruited using convenience sampling in public community spaces located within approximately 5 kilometers of Dhulikhel Hospital, including walking paths and roadside shops. Recruitment was conducted outside of hospital grounds, and no current hospital patients were approached.

The English-speaking study facilitator initially approached individuals to determine whether they appeared to understand English. If the individual did not demonstrate English comprehension, the accompanying Nepali-speaking collaborator explained the study in Nepali, confirmed interest, and obtained written informed consent. Both team members were present during all recruitment encounters. Individuals were eligible if they were ≥18 years old, primarily Nepali-speaking, and able to provide informed consent. No exclusions were made on the basis of education level, literacy, technology familiarity, or prior healthcare use.

The protocol specified a target sample of 20–30 participants for this exploratory feasibility study. No formal sample-size calculation was performed because the study was descriptive and was not designed for hypothesis testing; 30 participants were ultimately enrolled.

### Translation Procedure

Following informed consent, all conversational interactions were conducted between the English-speaking facilitator and the Nepali-speaking participant using ChatGPT-4o’s voice-to-voice translation system. The Nepali-speaking collaborator did not participate in the conversation itself. Before initiating the translation prompt, the facilitator began an audio recording using a separate device. The recording captured the entirety of the conversation, starting immediately before the translation prompt was given and ending after the participant’s final response to the last interview question.

Interviews were conducted using OpenAI’s paid ChatGPT service with the GPT-4o model on an iPhone 13 Pro running iOS 16.6.1 over the Ncell mobile network. A new ChatGPT session was initiated for each participant. The same standardized initialization prompt was used at the start of every interview to ensure procedural consistency:

“You are acting as a real-time translator between English and Nepali. I will first speak in English, and you will translate my words into Nepali. Then, a Nepali speaker will respond, and you will translate their response into English. Only translate what is said—do not add or summarize anything. Speak naturally and clearly. Begin now.”

After the prompt was given, the facilitator asked the following conversational questions in English, finalized after pre-testing:

1. “How old are you?”
2. “Do you go to Dhulikhel Hospital?”

If yes:
– “What did you go for?”
– “How did you get there?”
If no → proceed
3. “What do you do to stay healthy?”
4. “What is your favorite festival?”

– “How do you celebrate it?”

At the conclusion of the final question, the facilitator notified the participant that the interview had ended and stopped the recording. Audio files were transferred to secure, access-restricted cloud storage. These recordings were later reviewed by a bilingual evaluator, alongside the ChatGPT text transcripts.

Interview questions were intentionally non-diagnostic and low-risk, designed to elicit natural conversational speech rather than assess clinical decision-making. Prompts included demographic, health-adjacent, and culturally grounded topics to capture a range of structured and open-ended responses while minimizing potential harm from translation errors. This approach allowed evaluation of real-time translation fidelity under conditions representative of community health interactions without introducing clinical risk.

### Qualitative Coding Framework

We used an inductive, data-driven approach to characterize translation errors in ChatGPT-mediated Nepali–English interpretation. A single reviewer (American Council on the Teaching of Foreign Languages Advanced-Mid proficiency) reviewed each translated utterance and assigned one or more error tags based on observable deviations in meaning, tone, or phrasing from the speaker’s intended message. The coding process was iterative: categories were refined as additional transcripts were reviewed, and tag definitions were adjusted to ensure internal consistency across the dataset. After the final tag set was established, the reviewer conducted a second complete pass of all exchanges to ensure that each utterance was evaluated using the finalized and consistently applied coding framework. Because the aim was to describe the types of errors that occurred rather than to test or validate an existing taxonomy, coding decisions reflected the reviewer’s linguistic judgment rather than a predefined scheme. Common English loanwords routinely used by Nepali speakers, such as “hospital,” were not considered translation errors. Only one reviewer performed the coding, and therefore inter-rater reliability was not assessed.

Each translated utterance was rated on a 3-point scale to assess translation accuracy. A rating of 3 signifies high accuracy with the core meaning fully preserved, and the translation produced is natural sounding and requires no additional clarification. Minor differences in word order or slight awkwardness are acceptable as long as they do not confuse the listener or alter the intended meaning. A rating of 2 signifies moderate accuracy with the main idea preserved with some noticeable errors in grammar, word choice, or phrasing, while the participant’s intent is still generally understandable. There may be 1–2 moderate issues that require clarification, but the participant’s intent is still generally understandable. A rating of 1 signifies low accuracy, with the meaning significantly altered, unclear, or lost, causing breakdown in comprehension or communication.

Because individual translation outputs could deviate from the speaker’s intended message in more than one way, each translated utterance was eligible to receive multiple error tags. The final coding framework and corresponding examples are presented below.

**Distortion** refers to translations in which the meaning of the original sentence was changed, resulting in the listener receiving an incorrect interpretation of what the speaker intended. Example: “I go walking” translated as “I go by bus.”

**Too Formal/Stiff** was used when the translation was technically accurate but used vocabulary or sentence structure that felt bookish, formal, or overly polite compared to everyday conversational Nepali. This resulted in a tone that was socially distant or unnatural in context. Example: “सामान्यतः” (samanyata) for “usually,” instead of the more conversational “प्राय:” (praya).

**Omitted** refers to translation outputs in which a word or detail was left out, but the core meaning of the speaker’s message was still preserved. Example: “I go walking every morning” translated as “I go walking.”

**Over-interpretation** refers to cases where the translation added meaning that the speaker did not express, often by making a general statement more specific or implying intention or frequency not present in the original utterance. Example: “I go” translated as “I go often.”

**Participant Used English** was coded when the participant themselves switched to English, resulting in no translation being necessary for that utterance. This code does not reflect a system error, but rather a conversational choice by the speaker.

**Addition** was coded when the translation included extra descriptive words or clarifying detail that was not present in the original utterance, but did not change the speaker’s core meaning or intent. These additions made the translation longer or more explicit, but the listener would still understand the same message. Addition differs from Overinterpretation, in which the added content alters or extends the meaning beyond what the speaker intended. Example: “I go” translated as “I go there.”

**Clarification** was coded when the translation had to be repeated or rephrased because the initial translation attempt was unclear or not understood. Example: The speaker appears confused or asks the interviewer to repeat the statement.

**Grammar Error** refers to translations that were linguistically incorrect or unnatural in either Nepali or English. These errors did not necessarily change the overall meaning of the utterance, but they made the output sound awkward, unclear, or difficult to follow, affecting conversational ease and comprehension. Example: Incorrect verb agreement in Nepali or unnatural tense choice in English.

**Hindi Used** refers to cases where the translation incorporated Hindi words or phrasing in place of Nepali equivalents. While intelligible to many Nepali speakers, these substitutions shift the tone and cultural resonance of the utterance and can make the translation sound less natural or locally grounded. Example: Using “अच्छा” (achha) instead of “ठिक छ” (thik cha).

**Delay >3 Seconds** was coded when there was a clear pause of more than approximately three seconds between the participant’s speech and the system’s translation. These pauses interrupted the turn-taking rhythm of conversation and sometimes prompted the participant to repeat or modify their statement. Example: The participant completes a sentence, and the system waits ∼5 seconds before responding.

**ChatGPT Used English** was coded when the translation system responded in English instead of Nepali, despite the conversational context calling for Nepali output. Unlike Participant Used English, this reflects a system-level language selection issue, which may interrupt understanding for listeners who expected Nepali responses.

**No Translation Produced** was used when the system failed to return any translated output, even though the participant had spoken. This includes cases where the speech was not captured, the transcription failed, or the model returned an empty response. These events did not reflect meaning changes, but rather technical breakdowns in the translation process. Example: The participant speaks a full sentence and the system remains silent.

**Meaning Loss** refers to cases where the translation captured the general topic of the utterance but failed to carry over an important nuance, tone, or implied meaning. The listener would understand the broad idea, but not the emotional, cultural, or interpersonal intent the speaker conveyed. Example: A response expressing frustration is translated neutrally, reducing emotional meaning.

**Interrupted/Cutoff** referred to translations where the output terminated prematurely, leaving the sentence incomplete or the message unresolved. These cases resulted in the listener receiving only a partial utterance, requiring repetition or repair. Example: “I have had pain for two weeks in my hand” → “I have had pain for two weeks—”.

**Too Casual/Informal** was coded when the translation reduced or removed politeness, introduced slang, or dropped honorific language in a way that altered how the speaker’s social tone was conveyed. This included cases in which respectful Nepali forms (e.g., tapai, hajur) were replaced with familiar or lower-register forms (e.g., timi, ta). While these shifts did not necessarily change the core meaning, they resulted in the speaker sounding disrespectful, overly familiar, or inconsistent with Nepali conversational norms. Informal wording was not coded when it appropriately reflected the speaker’s original tone. Example: Tapai (“you,” respectful) translated as timi (“you,” casual).

**Number Error** refers to translation output in which quantitative information (such as age, duration, or frequency) was mis-transcribed or misinterpreted. These errors are notable because even small numeric inaccuracies can change the factual meaning of the message. Example: The speaker reports their age as “67” and ChatGPT translated it as “47.”

**Nepali Dialect** was coded when the translation used a form of Nepali that was linguistically correct but not the appropriate dialect for the region, setting, or social context of the conversation. While still understandable, the dialect choice could shift tone, familiarity, or cultural alignment, making the utterance sound out of place or unnatural in the interaction. Example: “Hamra” instead of the standard “Hamro.”

**Wrong Language** was coded when the translation was produced in English instead of Nepali (or vice-versa), resulting in a disruption of the intended language exchange. This occurred even when the semantic content was correct, and the issue primarily affected conversational flow and accessibility. Example: A Nepali response prompt yields an English output.

### Ethics Statement

The protocol was reviewed and approved by the Institutional Review Committee of Kathmandu University School of Medical Sciences (IRC-KUSMS; approval no. 171/25) for implementation on 16 July 2025. All participants provided written informed consent and separately authorized audio recording after the study procedures and privacy risks were explained in Nepali by a bilingual translator approved by Dhulikhel Hospital-Kathmandu University Hospital. Participation was voluntary, and participants could skip questions or withdraw at any time.

Potential risks included loss of privacy, social discomfort, and inconvenience. Participants were asked not to disclose identifying information and were informed that their speech would be processed by ChatGPT on external servers. Names were collected only on consent forms and were not linked to study data. Recordings were stored securely with access restricted to the approved research team. Accidentally disclosed identifiers were removed from transcripts, and only de-identified data were used in the analysis and publication.

## Results

### Translation accuracy by direction

Across 485 translated utterances, 282 (58.1%) received the highest accuracy rating of 3, indicating preservation of core meaning with natural-sounding communication; 134 (27.6%) received a rating of 2, and 69 (14.2%) received a rating of 1 (Table 1). English-to-Nepali (E→N) translations had a mean accuracy rating of 2.63 ± 0.53, compared with 2.23 ± 0.86 for Nepali-to-English (N→E) translations.

**Table 1.**
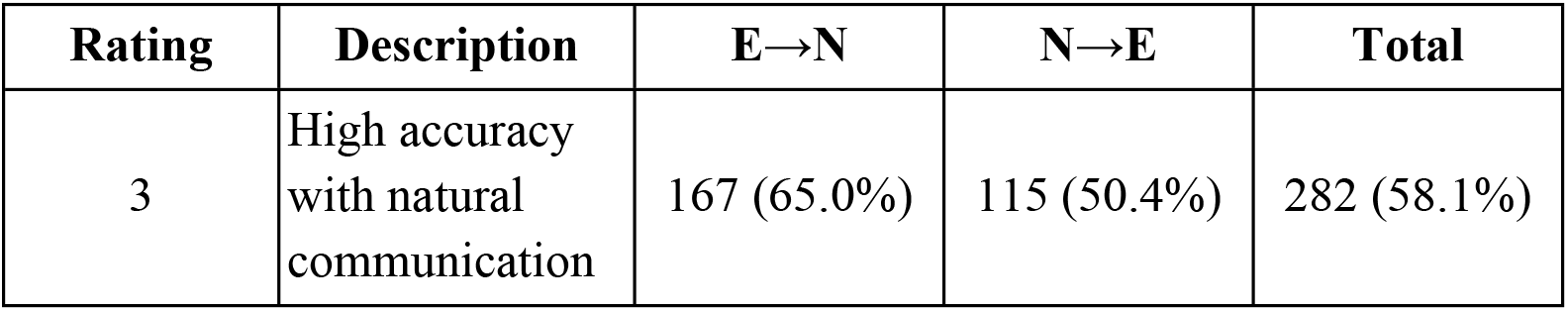
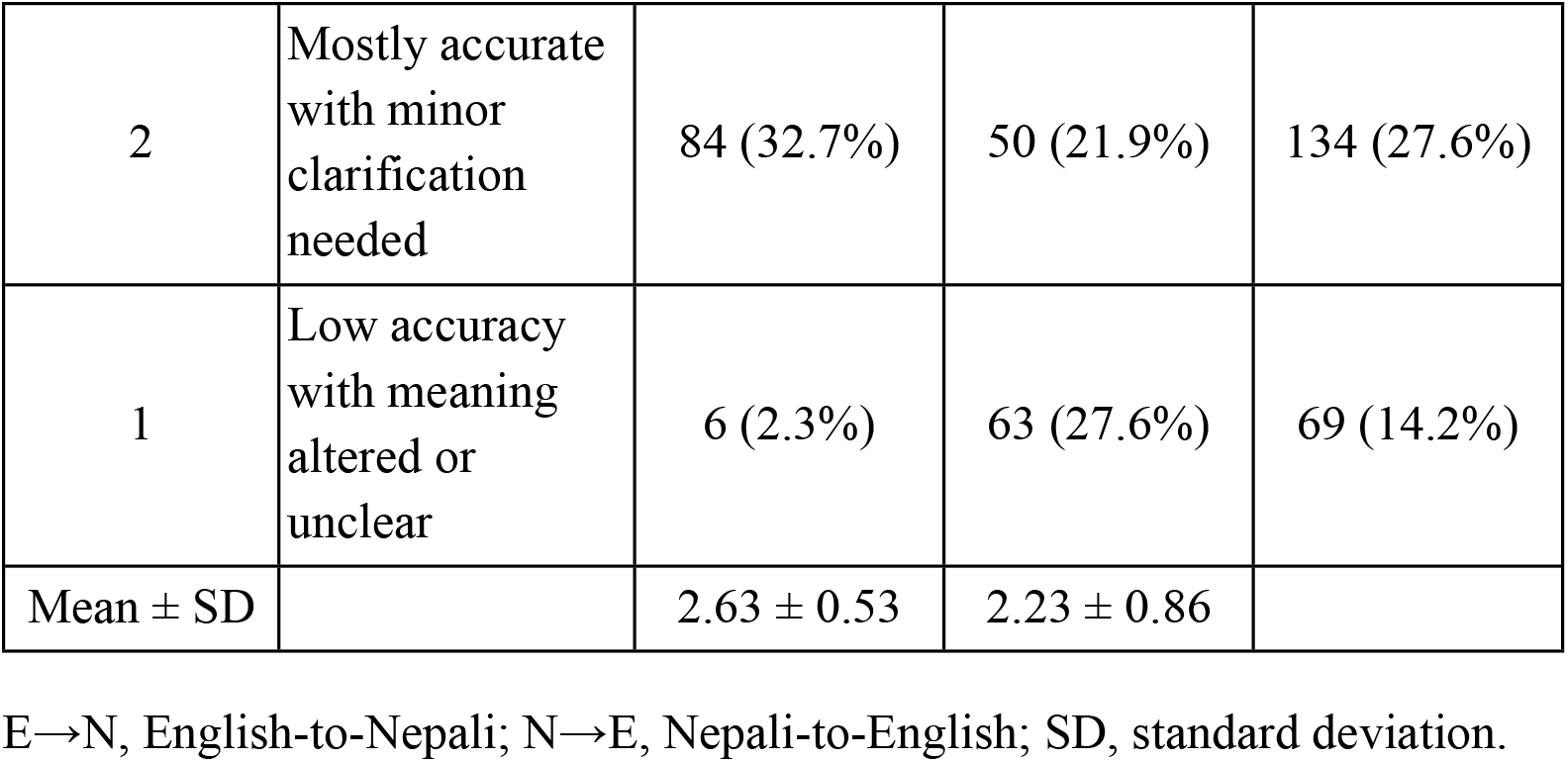
Distribution of Translation Accuracy Ratings by Direction (English→Nepali and Nepali→English) The distribution of accuracy ratings differed by translation direction (Fig 1). Among E→N translations, 167 of 257 (65.0%) received a rating of 3 and 6 (2.3%) received a rating of 1. Among N→E translations, 115 of 228 (50.4%) received a rating of 3 and 63 (27.6%) received a rating of 1. Of the 69 low-accuracy translations, 63 (91.3%) occurred in the N→E direction.

**Fig 1.**
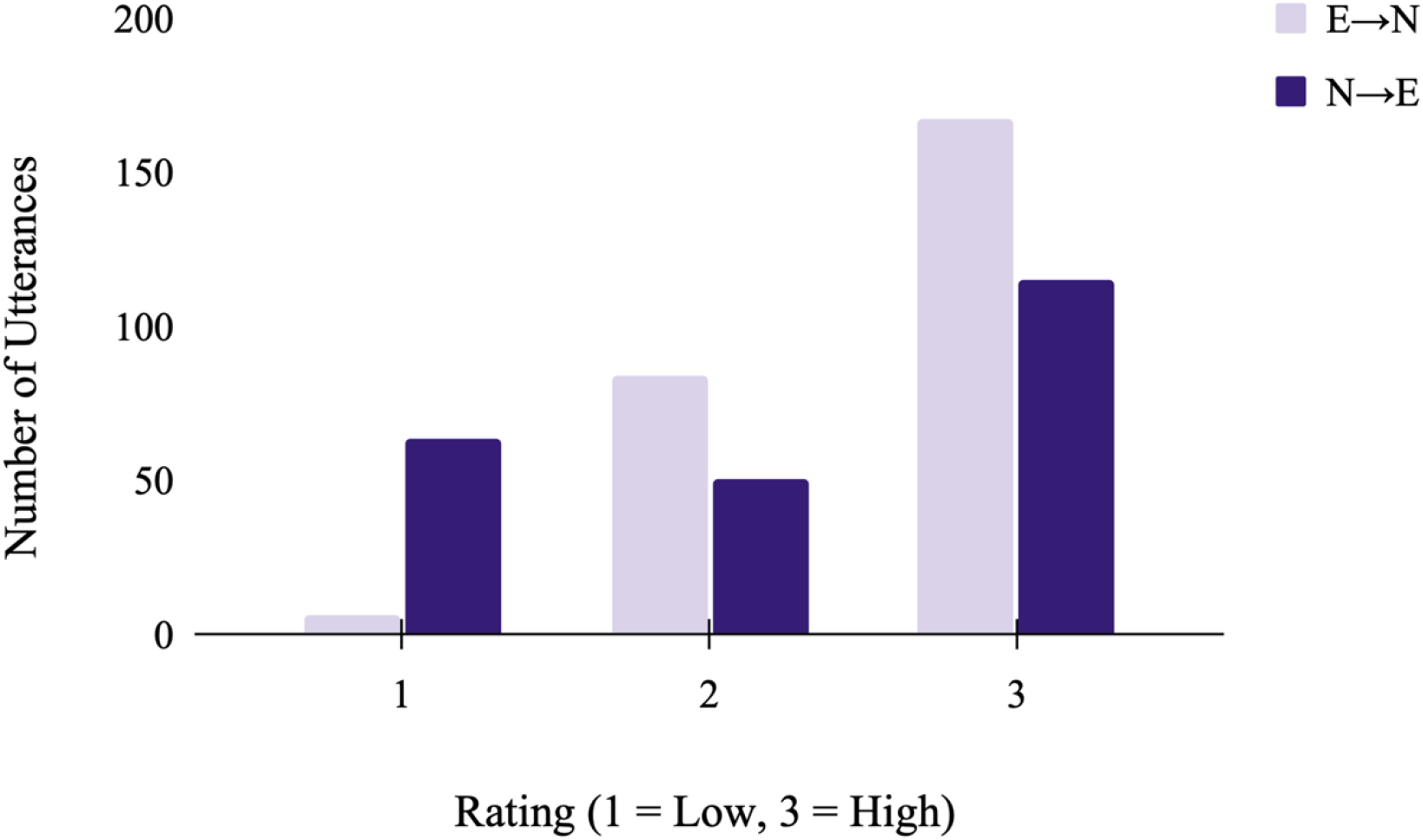
Distribution of translation accuracy ratings by translation direction. Bar heights reflect the number of translated utterances receiving each accuracy rating (1 = low accuracy, 2 = moderate accuracy, 3 = high accuracy). English-to-Nepali (E→N) translations had a greater proportion of high-accuracy ratings, whereas Nepali-to-English (N→E) translations had a greater proportion of low-accuracy ratings.

### Accuracy by interview question

Translation accuracy varied across interview questions (Table 2). The highest mean E→N accuracy rating was observed for the age question (Q0; 2.79 ± 0.48), followed by questions regarding Dhulikhel Hospital use and associated follow-up questions (Q1; 2.68 ± 0.47) and festival-related questions (Q3; 2.64 ± 0.48). The lowest E→N mean rating occurred for the question “What do you do to stay healthy?” (Q2; 2.33 ± 0.63).

**Table 2.** Translation quality scores by interview question.

| Question ID | Description | E→N Mean ± SD | n (E→N) | N→E Mean ± SD | n (N→E) |
| --- | --- | --- | --- | --- | --- |
| **Q0** | Age Question<br>("How old are you?") | $2.79 \pm 0.48$ | 33 | $2.26 \pm 0.96$ | 34 |
| **Q1** | "Do you go to Dhulikhel Hospital?"<br>(including any follow-up questions asked only if 'yes') | $2.68 \pm 0.47$ | 90 | $2.33 \pm 0.83$ | 92 |
| **Q2** | "What do you do to stay healthy?" | $2.33 \pm 0.63$ | 36 | $1.85 \pm 0.86$ | 34 |
| **Q3** | "What is your favorite festival?"<br>(and if answered:<br>"How do you celebrate it?") | $2.64 \pm 0.48$ | 66 | $2.26 \pm 0.80$ | 68 |
| **Other** | Instructions, conclusions, small talk | $2.63 \pm 0.61$ | 32 | -- | 0 |

E→N, English-to-Nepali; N→E, Nepali-to-English; SD, standard deviation. Follow-up questions are included under Q1 and Q3 because translation accuracy was scored at the question-group level.

For N→E translations, mean ratings were 2.26 ± 0.96 for Q0, 2.33 ± 0.83 for Q1, and 2.26 ± 0.80 for Q3. The lowest mean N→E rating was observed for Q2 (1.85 ± 0.86). Across all question categories evaluated in both directions, standard deviations were larger for N→E than E→N translations.

### Coded translation and conversational deviations

A total of 19 distinct deviation types were identified using the inductively developed coding framework (Table 3). Because individual utterances could receive more than one code, percentages represent the proportion of all assigned coding tags rather than the proportion of translated utterances.

**Table 3.**
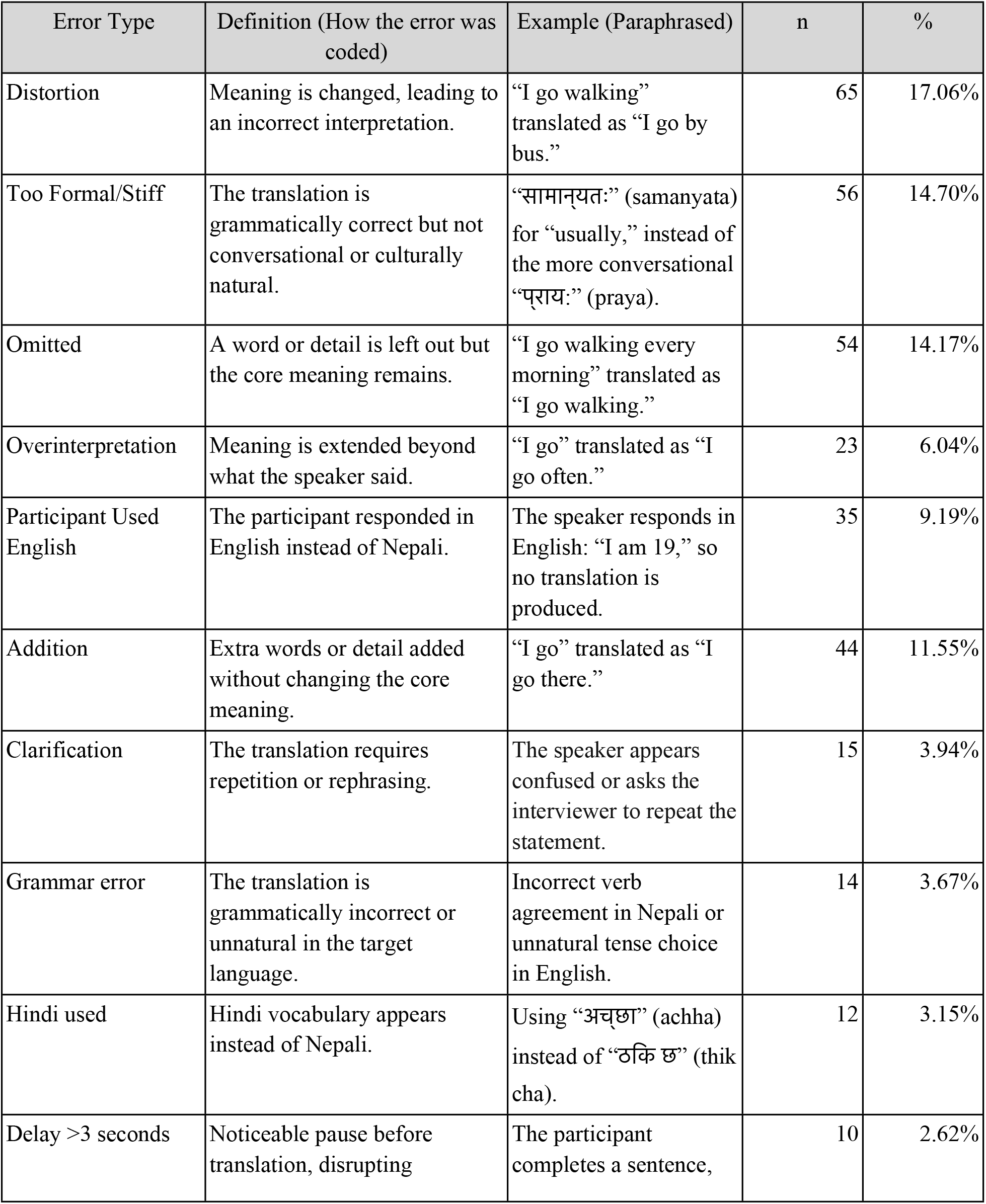
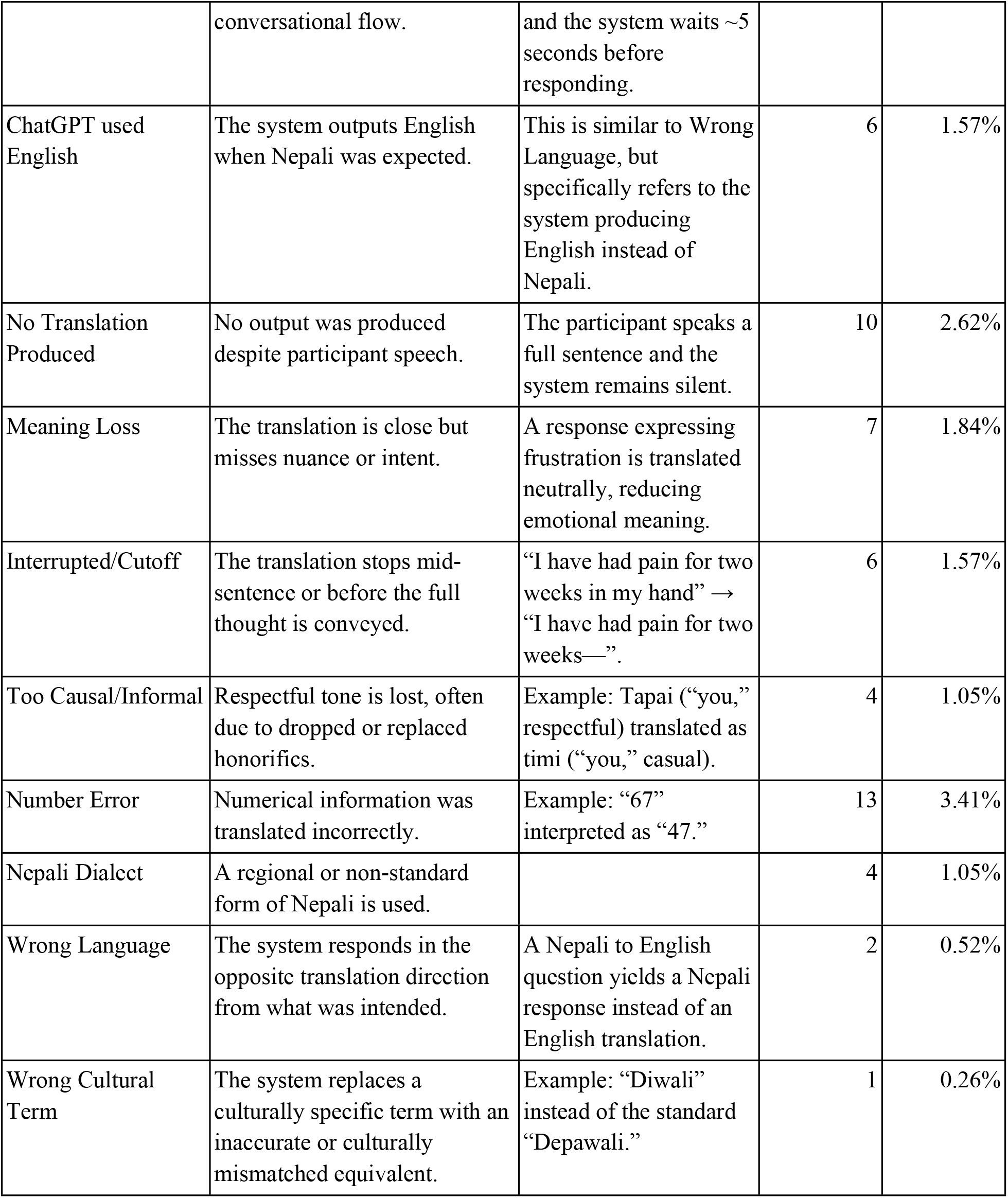
Frequency, definitions, and paraphrased examples of translation errors identified across English–Nepali conversational exchanges.

Distortion was the most frequently assigned code (n = 65, 17.1%), followed by Too Formal/Stiff phrasing (n = 56, 14.7%), Omission (n = 54, 14.2%), and Addition (n = 44, 11.5%). Overinterpretation was coded in 23 instances (6.0%). Participant Used English, which reflected participant language choice rather than a system translation error, accounted for 35 coded instances (9.2%).

Less frequent coded deviations included Clarification (n = 15, 3.9%), Grammar Error (n = 14, 3.7%), Number Error (n = 13, 3.4%), Hindi Used (n = 12, 3.2%), Delay >3 Seconds (n = 10, 2.6%), and No Translation Produced (n = 10, 2.6%). Meaning Loss was coded seven times (1.8%). ChatGPT Used English and Interrupted/Cutoff were each coded six times (1.6%). The remaining coded deviations each accounted for 1.1% or less of assigned tags.

### Representative translation examples

Representative examples across accuracy ratings are presented in Table 4. Translations rated 3 preserved the speaker’s intended meaning despite minor differences in word order, repetition, or contextual detail. For example, a participant’s repeated statement of being “twenty two, twenty two years” retained the same factual meaning in the translated output. In another exchange, the model appropriately translated the idiomatic expression sathi bhai as “friends,” rather than rendering its individual components literally as “friends and brothers.”

**Table 4.**
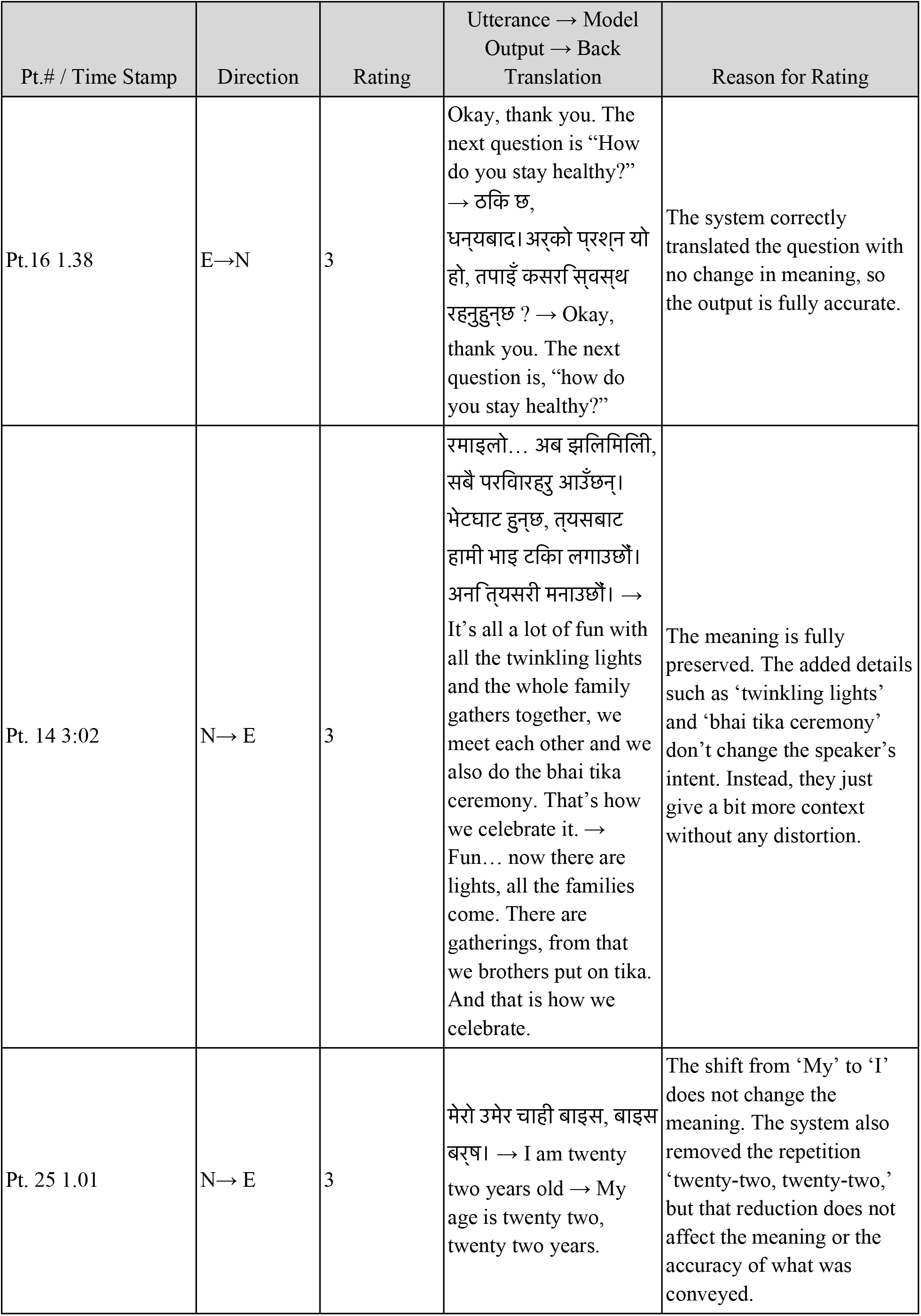
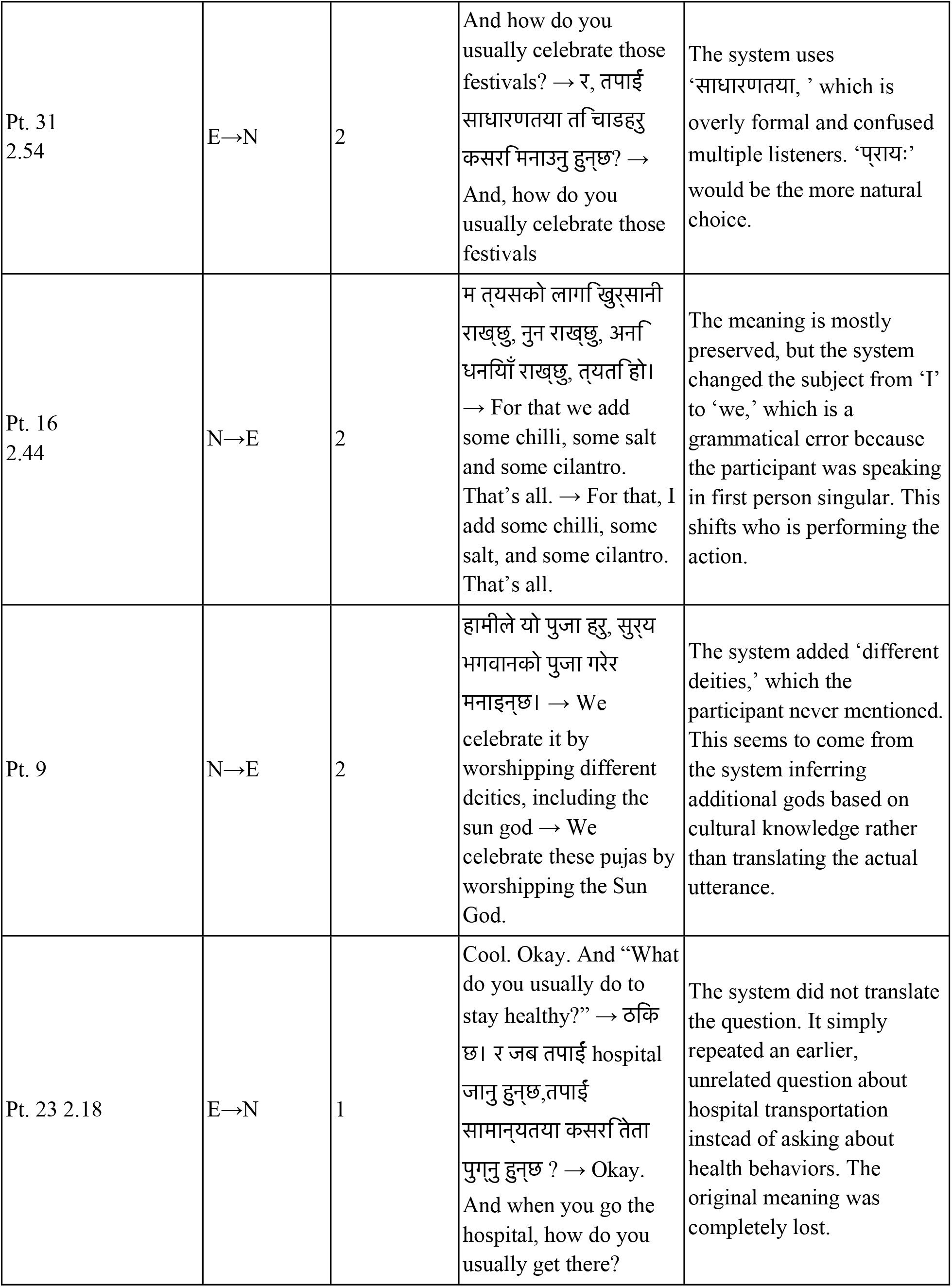
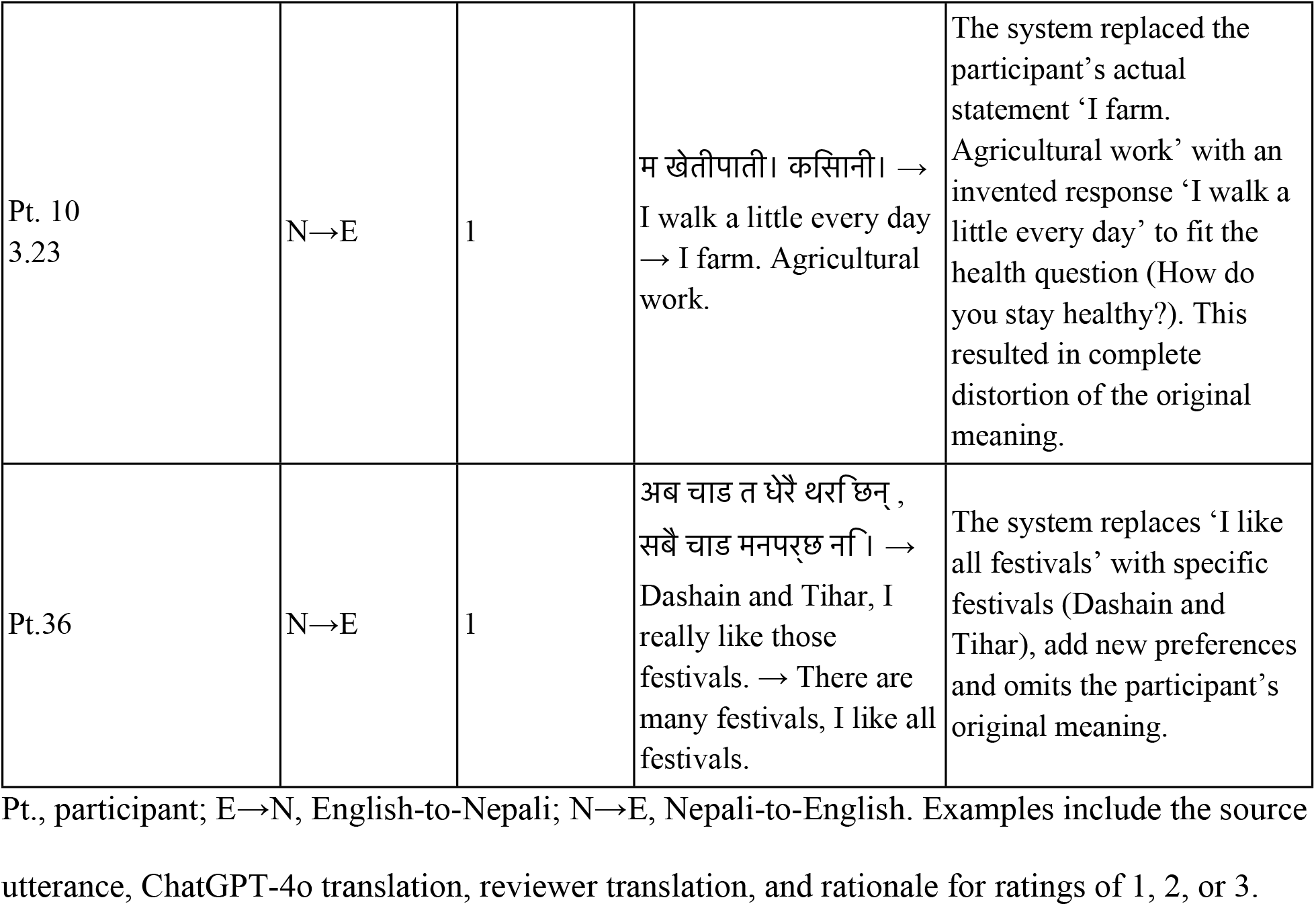
Representative English–Nepali translations across accuracy ratings.

Translations rated 2 generally preserved the main message but included changes in grammar, phrasing, or specificity. In one N→E translation, a first-person statement describing the addition of chili, salt, and cilantro was translated using “we” rather than “I.” In another, the system added “different deities” to a statement describing worship of the Sun God.

Translations rated 1 substantially altered or failed to preserve the original meaning. When the facilitator asked, “What do you usually do to stay healthy?”, the system instead produced an earlier question about transportation to the hospital. In another exchange, the participant’s statement “I farm. Agricultural work” was translated as “I walk a little every day.” A participant’s statement that they liked all festivals was also translated as a specific preference for Dashain and Tihar.

## Discussion

The present study evaluated ChatGPT-4o’s voice-to-voice translation capability during field-based conversations in rural Nepal, as assessed by a bilingual Nepali–English reviewer. Most translations preserved the speaker’s core intended meaning, suggesting that ChatGPT-4o may facilitate real-time Nepali–English communication in some low-stakes settings. However, translation performance differed substantially by direction, with English-to-Nepali translations demonstrating greater accuracy and consistency than Nepali-to-English translations. The system also produced errors that altered intended meaning or introduced content that had not been spoken. These findings are particularly relevant to research and healthcare contexts, where fluent but inaccurate translations may be accepted as reliable, misrepresent participant responses, or introduce misinformation. Human oversight and mechanisms for verification therefore remain important when AI-mediated translation is used in settings where translation accuracy has meaningful consequences.

Translation errors varied substantially in their potential effect on communication. The most frequently identified errors included distortion of intended meaning, overly formal or unnatural phrasing, and omission of words or details. Some primarily affected conversational naturalness. For example, ChatGPT-4o translated “usually” using “सामान्यतः” (samanyata) rather than the more conversational “प्राय:” (*praya*), producing wording that was understandable but less natural in spoken Nepali. The model also sometimes shifted between honorific and non-honorific forms, preserving semantic meaning while altering the level of respect conveyed. Conversely, it appropriately translated the idiomatic expression sathi bhai as “friends,” rather than rendering it literally as “friends and brothers.” Other errors changed the information conveyed to the listener. Low-accuracy translations occurred predominantly in the Nepali-to-English direction, accounting for 63 of the 69 translations rated as 1. This directional imbalance raises concern that the increasing accessibility of generative AI translation may not be accompanied by equivalent reliability across languages or translation directions. This issue may be particularly relevant for lower-resource languages such as Nepali, for which prior studies have identified persistent limitations in machine translation performance related to language-resource availability and linguistic and cultural features of the source language [13,19,22]. However, the directional difference observed in the present study cannot be attributed to these factors alone. English prompts were standardized and repeated across interviews, whereas Nepali responses were spontaneous and varied in length, clarity, and complexity. Hesitant or quiet speech, background noise, overlapping conversation, and participant unfamiliarity with the tool may also have affected speech recognition before translation occurred. These differences could have contributed to lower and more variable Nepali-to-English accuracy and represent important alternative explanations for the observed directional asymmetry.

To our knowledge, this is the first study to evaluate real-time, voice-to-voice English–Nepali translation using ChatGPT-4o. Although differences in language pairs and study design limit direct comparison, the findings are consistent with prior research suggesting that machine translation may facilitate selected forms of communication while remaining unreliable for higher-stakes use. Hudelson and Chappuis found that clinicians frequently achieved consultation goals using voice-to-voice translation despite lower satisfaction with communication quality [5]. Similarly, Kong et al. reported high sentence-level accuracy for ChatGPT-generated discharge translations, while many complete instruction sets contained at least one error [11]. The present study extends this literature by evaluating bidirectional, real-time voice translation during field-based community conversations rather than controlled written translation or structured clinical encounters.

The error patterns observed in this study further support the distinction between translation fluency and translation fidelity. Prior comparisons of generative AI translation with conventional machine translation and professional human translators have shown that fluent output may still contain additions, omissions, and context-dependent distortions [11,21–23]. A similar pattern was observed in the present study, where some translations were coherent and natural despite changing the speaker’s intended meaning or introducing information that had not been expressed. Reports of speech-processing systems generating words or complete sentences absent from the original audio further reinforce concerns that plausible output may be interpreted as accurate when it is not [24]. This distinction is especially important in research and healthcare, where translated statements may be used to characterize participant experiences, identify community needs, or inform clinical decisions. Strong aggregate translation performance should therefore not be interpreted as sufficient evidence of reliability for unsupervised use when individual translation errors may carry substantial consequences [4,11,25]. These findings suggest that aggregate accuracy alone may not adequately capture the risks of AI-mediated translation, particularly when individual errors are difficult to detect and substantially alter meaning.

Field observations provided additional context regarding the conditions surrounding AI-mediated communication. In group settings, some participants looked to others for cues before answering, and participants sometimes appeared more comfortable after observing the translation process. Community interest in the tool was generally positive, with some participants independently identifying potential applications for communicating with non-Nepali-speaking individuals. Although these observations were not systematically collected or formally analyzed and should not be interpreted as evidence of acceptability or user behavior, they highlight questions that warrant further study. Future evaluations should also assess community perspectives on AI-mediated translation, including existing patterns of use, trust in AI-generated outputs, and willingness to rely on these tools for communication in real-world settings.

### Strengths and Limitations

This study has several strengths. ChatGPT-4o was evaluated during live, bidirectional community conversations, allowing translation performance to be assessed under field conditions rather than using standardized written text alone. The same English-speaking facilitator, device, account, model version, initialization prompt, and core questions were used across encounters, providing consistency in the translation procedure. Translation quality was also evaluated using both an overall accuracy rating and an inductively developed, meaning-focused error taxonomy, allowing errors with different implications for conversational naturalness and fidelity to be characterized separately. The inclusion of spontaneous Nepali responses further increased the practical relevance of the evaluation.

Several limitations should be considered when interpreting the findings. Participants were recruited through convenience sampling from a single geographic area, limiting generalizability to other regions, dialects, populations, and clinical settings. Although age, sex, and literacy were collected, these characteristics were not incorporated into the analysis, preventing assessment of whether participant characteristics influenced technology use or translation performance. Accuracy ratings and error coding were completed by a single bilingual reviewer, therefore inter-rater reliability was not assessed; subjective classification may therefore have influenced both accuracy scores and error categorization. Differences between translation directions were also potentially confounded by the use of standardized, repeated English prompts compared with spontaneous Nepali responses. Uncontrolled differences in speech volume, hesitation, background noise, overlapping conversation, and familiarity with the technology may have biased Nepali-to-English performance toward lower accuracy independently of translation direction. In addition, the study could not distinguish errors introduced during speech recognition from those arising during translation, did not compare ChatGPT-4o with professional interpreters or alternative translation systems, and evaluated only one initialization prompt and model configuration. Finally, generative AI systems are rapidly evolving, and newer model versions have become available since data collection. These findings therefore represent the performance of a specific system and configuration at a defined point in time and may not generalize to current or future versions.

## Conclusion

This study demonstrates both the potential and current limitations of real-time, AI-mediated voice translation for Nepali–English communication. ChatGPT-4o frequently preserved intended meaning during field-based conversations, but translation accuracy was asymmetric and occasional fluent outputs substantially altered or introduced information not expressed by the speaker. These findings are particularly important as multimodal models capable of real-time audio interaction become increasingly accessible. Although such tools may help bridge language barriers in selected low-stakes settings, the present findings do not support unsupervised use where translation errors could influence research findings or healthcare decisions. Future evaluations should assess newer models and alternative translation systems across diverse lower-resource languages and real-world conversational conditions, particularly as AI translation capabilities continue to evolve rapidly. Research should also evaluate the effectiveness of safeguards for identifying, flagging, and verifying potentially inaccurate outputs before these systems are implemented in high-consequence settings.

## Data Availability

All de-identified data underlying the translation accuracy and error-classification findings reported in this article are provided in S1 Dataset. Participant identifiers in S1 Dataset are replacement study codes and do not correspond to the identifiers used on participant consent forms. Audio recordings and full conversation transcripts are not publicly available because they contain potentially identifying human-participant conversational data, and participant consent did not authorize unrestricted public release of these materials. In accordance with the approved study protocol, these restricted materials will be retained only through July 16, 2027, and will then be securely destroyed. Until that date, qualified researchers may request access to de-identified transcript data by contacting the Institutional Review Committee, Kathmandu University School of Medical Sciences (IRC-KUSMS), at. Requests will be considered for research purposes subject to applicable institutional review and approval by Dhulikhel Hospital-Kathmandu University Hospital.

https://docs.google.com/spreadsheets/d/1KSL_DF2iQ4Ez5lP81kkpmPaARVA8UQ5OlQrEtjNNFi4/edit?usp=sharing

## Acknowledgements

We thank the community members who participated in this study for their time and willingness to engage with an unfamiliar AI-mediated translation tool. We are grateful to Silvia Pradhan, a community contributor and graduate of Purbanchal University, for transcribing the Nepali-language conversations used in this analysis. We also thank Dhulikhel Hospital and its founder, Prof. Dr. Ram Kantha Makaju Shrestha, for facilitating the partnerships and institutional support that enabled this field study to be established and conducted within a limited timeframe. We further acknowledge the University of Washington Global Health Immersion Program for providing ethical guidance, facilitating institutional partnerships, and supporting the development and implementation of the project.

## Supporting Information

**S1 File.** De-identified dataset of ChatGPT-4o English–Nepali translation accuracy ratings and error classifications, including utterance-level accuracy scores and coded translation or conversational deviations.

